# A phase I study of oral vitamin D3 in boys with X-linked adrenoleukodystrophy

**DOI:** 10.1101/2021.12.28.21267861

**Authors:** Keith Van Haren, Kristen Cunanan, Avni Awani, Meng Gu, Dalia Peña, Lindsay C. Chromik, Michal Považan, Nicole C. Rossi, Jennifer Winterbottom, Jordan Goodman, Vandana Sundaram, Gerald V Raymond, Tina Cowan, Gregory M. Enns, Emmanuelle Waubant, Lawrence Steinman, Peter B. Barker, Daniel Spielman, Ali Fatemi

**Affiliations:** Department of Neurology, Stanford University School of Medicine, Palo Alto, CA; Department of Pediatrics, Stanford University School of Medicine, Palo Alto, CA; Quantitative Sciences Unit, Stanford University School of Medicine, Palo Alto, CA; Department of Radiology, Stanford University School of Medicine Palo Alto, CA; Russell H. Morgan Department of Radiology and Radiological Science, The Johns Hopkins University School of Medicine, Baltimore, MD; The Kennedy Krieger Institute, Baltimore, MD; Department of Genetic Medicine, The Johns Hopkins University School of Medicine, Baltimore, MD; Department of Pathology, Stanford University School of Medicine, Palo Alto, CA; Department of Neurology, University of California at San Francisco, San Francisco, CA; Department of Neurology, The Johns Hopkins University School of Medicine, Baltimore, MD

## Abstract

**Objective:** Vitamin D status has been linked to risk of inflammatory brain lesions. We sought to assess the safety and pharmacokinetics of oral vitamin D dosing regimens in boys with X-linked adrenoleukodystrophy (ALD).

**Methods:** In this open-label, multi-center, phase I study, we enrolled 21 ALD males without brain lesions, aged 1.5 to 25 years to oral vitamin D supplementation for 12 months. Our primary outcome was attainment of plasma 25-hydroxyvitamin D levels in target range (40-80ng/ml) at 6 and 12 months. Secondary outcomes included safety and glutathione levels in brain and blood. Participants were initially assigned to a fixed dosing regimen starting at 2,000 IU daily, regardless of weight. Following a mid-study safety assessment, we modified the dosing regimen so all subsequent participants were assigned to a weight-stratified dosing regimen starting as low as 1,000 IU daily.

**Results:** Between October 2016 and June 2019, we recruited 21 participants (n=12 fixed dose; n=9 weight-stratified) with a median age and weight of 6.7 years and 20 kilograms. Most participants achieved target plasma vitamin D levels at 6 and 12 months regardless of dosing regimen. In the fixed dose regimen, 6 of 12 participants had asymptomatic elevation in urine calcium:creatinine or plasma 25-hydroxyvitamin D; no laboratory deviations occurred with the weight-stratified regimen. Glutathione levels increased between baseline and 12 months in the brain but not in the blood.

**Conclusions:** Our weight-stratified vitamin D dosing regimen was well-tolerated and achieved target 25-hydroxyvitamin D levels in most participants. Brain glutathione levels increased over the 12-month trial period.

**Clinicaltrials.gov identifier:** NCT02595489

**Classification of Evidence:** This study provides Class II evidence that a weight-stratified dosing regimen of vitamin D supplementation is safe, well-tolerated, and effective at achieving moderately high vitamin D levels in boys with ALD.

**Sources of funding:** NIH/NINDS K23NS087151

## INTRODUCTION

X-linked adrenoleukodystrophy (ALD) is a panethnic monogenic disorder with an incidence of 1-2 per 20,000 births^1-3^. ALD is caused by loss of function mutations in a gene (*ABCD1*) encoding a peroxisomal surface protein that facilitates metabolism of very long chain fatty acids. Approximately 2/3rds of males with ALD develop an inflammatory cerebral demyelinating phenotype (cerebral ALD) over their lifetime; females are rarely affected^4, 5^. The distribution of cerebral ALD lesion formation varies by age. Although cerebral ALD lesions can appear at across the lifespan, approximately half of lesions manifest in the first 10 years of life^4, 5^. Most lesions originate in the occipital or frontal white matter^6, 7^. Cerebral ALD lesions share histologic similarities with multiple sclerosis but do not respond to multiple sclerosis therapies and typically enlarge relentlessly unless hematopoietic stem cell transplantation is initiated while the lesion is still small^8, 9^. As ALD is added to a growing number of newborn screening panels worldwide, a new era of surveillance for early signs of cerebral ALD lesions that may facilitate preventive therapies^3, 10^.

Although no preventive therapies currently exist for the cerebral ALD phenotype, epidemiologic evidence from multiple sclerosis suggests that vitamin D insufficiency predisposes to the development of brain lesions while vitamin D supplementation may confer a reduction in brain lesions and inflammatory markers^11-17^. Although vitamin D’s immunoregulatory effects are well recognized, vitamin D has more recently been linked to a range of antioxidant effects, including augmentation of glutathione, an abundant and ubiquitous antioxidant whose deficit has been implicated in many diseases, including ALD^18-21^.

We undertook the current study primarily to assess the safety and biomarkers of various oral supplementation regimens capable of safely achieving moderately high levels (40-80ng/ml) of 25-hydroxyvitamin D in ALD boys. Secondarily, we sought to assess vitamin D’s potential role in augmenting glutathione levels in the brain and blood of ALD boys.

## METHODS

### Research question

The primary objective of our study was to identify a dosing regimen for safely achieving plasma 25-hydroxyvitamin D levels between 40-80ng/ml in boys with ALD. We also hypothesized that oral vitamin D supplementation would increase glutathione levels in the frontal and occipital white matter and whole blood.

### Standard protocol approvals, registrations, and participant consents

The trial was registered ClinicalTrials.gov (NCT02595489) prior to participant enrollment. The protocol and consents were approved by the institutional review boards at Stanford University and Kennedy Krieger Institute. All participants provided written informed consent.

### Inclusion, Exclusion, and Enrollment

Participants were eligible for enrollment if they were males with a molecular diagnosis of X-linked ALD between the age of 1.5 and 25 years, without evidence of gadolinium-enhancing cerebral demyelination, had baseline 25-hydroxyvitamin D level less than or equal to 60ng/ml, and had normal values for serum calcium, creatinine, phosphorus, parathyroid hormone, and urinary calcium:creatinine ratio. Participants were excluded if they had a history of liver or kidney disease, nephrolithiasis, hyperthyroidism, inflammatory bowel disease, medications interfering with intestinal absorption, or had contraindication to completing a brain MRI every six months. Participants were enrolled at two sites: Lucile Packard Children’s Hospital and Kennedy Krieger Institute.

### Trial design

This study was originally conceived as a single-arm dose-escalation study designed to assess two fixed daily doses of vitamin D (2,000 IU vs 4,000 IU). However, after interim safety analysis suggested higher rates of laboratory deviations (e.g. 25-hydroxyvitamin D > 80ng/ml) in younger participants, the study was modified to a weight-based protocol that stratified dosing regimens based on each participant’s bodyweight at time of enrollment.

Following enrollment, the principal study site mailed a 3-month supply of sublingually dissolvable 1,000 IU (25mcg) vitamin D3 tablets. Participants were instructed to administer the specified number of tablets once daily. Drug was manufactured by Continental Vitamin Company. One participant on the weight-stratified regimen was provided with 1,000 IU tablets from a separate manufacturer (Renzo’s Vitamins, Inc) to conform with a vegan diet.

The dosing regimens and conditional escalation parameters are summarized in **Figure 2**. Briefly, all participants in the fixed-dose group were assigned to an initial 6-month period at 2,000 IU daily. If the participant’s plasma 25-hydroxyvitamin D levels remained <60ng/ml at the 6-month study visit, the dose was increased to 4,000 IU daily. All participants in the weight-stratified dose group were assigned to either 1,000 IU, if baseline weight < 20 kg, or 2,000 IU, if baseline weight > 20 kg. If the participant’s plasma 25-hydroxyvitamin D levels remained <40ng/ml at the 6-month study visit, the dose was increased to either 2,000, 3,000, or 4,000 IU, depending on baseline weight. Participants who did not qualify for dose escalation remained at their starting dose. Pre-specified dose reduction parameters required dose reductions in all participants with plasma 25-hydroxyvitamin D levels exceeding 80ng/ml. To account for variations UV exposure in determining vitamin D levels, skin tone was scored on a scale of 1 (very fair) to 6 (black); sun exposure was assessed at baseline, 6, and 12 months using questionnaires.

### Primary outcome

The primary outcome is the proportion of participants who achieved a plasma 25-hydroxyvitamin D level in the target range (40-80ng/ml) at 6 months and 12 months without a triggered dose reduction. Vitamin D levels were measured by the clinical laboratories at participating institutions, both of which employed tandem mass spectroscopy. For participants missing a vitamin D level at any timepoint the most recent preceding vitamin D level was used in its place.

### Measurement of glutathione levels in brain

Single-voxel proton magnetic resonance spectroscopy (MRS) was attempted for all participants at baseline, 6 and 12 months. *In vivo* MRS data were acquired on either a GE or Philips 3T scanner using the MEGA-PRESS sequence^22, 23^. Occipital and frontal white matter regions were selected because they are the typical locations of cerebral ALD lesion formation^7^. For each subject, data were sequentially acquired from two 2×6×2cm voxels located centered in the occipital and frontal white matter and frontal lobes (**Figures 4A, 4B**). The GSH editing pulses for the MEGA-PRESS sequence were of 20 millisecond duration 180º Gaussian pulses applied at 4.56 and 7.5ppm. A total of 256 transients were acquired for each voxel using a 5000 Hz spectral bandwidth, 2048 data points, TE/TR=80ms/2s, and a total scan time of 8:40 minutes. GSH was quantified by fitting the 2.95 ppm peak in the edited spectrum and referenced to total creatine^23^. We centralized our spectroscopy analysis to reduce inter-site variability. A designated expert (MG) was responsible for assessing the quality of the acquired spectra. For each measure, criteria for inclusion in final analyses were: (1) adequate shimming with linewidth <18 Hz, (2) flat baseline, and (3) discernible GSH peak at 2.95 ppm in the edited spectrum.

### Measurement of glutathione levels in whole blood

Blood collection, preparation and analysis were carried out using a Sciex 4500 triple quadrupole mass spectrometer and Analyst software as previously described ^24^. In brief, blood samples were refrigerated immediately following collection and processed within 24 hours by adding a precipitating solution of sulfosalicylic acid containing the derivatizing agent N-ethylmaleimide (NEM). Derivatized samples were stored at -80°C prior to analysis. Samples were analyzed by LC-MS/MS using stable-isotope internal standards of GSH (GSH-^13^C,^15^N) and GSSG (GSSG-^13^C,^15^N) (Cambridge Isotope Laboratories, Inc, Tewksbury, MA) for quantitation. GSH-NEM and GSSG ions and fragments monitored in the positive mode using transitions *m/z* 433>304 and *m/z* 613>355, respectively. Stable-isotope internal standards were monitored as *m/z* 435>306 (GSH-^13^C, ^15^N-NEM) and *m/z* 617>359 (GSSG-^13^C,^15^N).

### Safety criteria and adverse events

Safety was assessed using clinical, laboratory, and radiologic measures. Participants were monitored for adverse clinical events through quarterly surveys to assess for signs and symptoms associated with hypercalcemia and general adverse events. We monitored for deviations in laboratory markers associated with excessive vitamin D intake: plasma 25-hydroxy vitamin D levels and calcium were measured quarterly while urine calcium:creatinine levels were assessed every 6 months. Brain MRIs were completed every 6 months to screen for the appearance of cerebral ALD lesions. Pre-specified events would trigger dose reduction of daily vitamin D supplementation; specified events included plasma 25-hydroxyvitamin D levels >80ng/ml and persistently elevated urine calcium:creatinine. If urine calcium:creatinine ratio was elevated at 6-month screening, it triggered three subsequent monthly measures with dose reduction implementation only if two of those three measures remained elevated (i.e. persistent elevation).

### Statistical analysis

All participants who received at least one dose of study drug are included in the safety analysis and in the primary analysis. Appropriate summary statistics are used to present demographic and primary outcome data, overall and by dosing regimen. We used Wilcoxon signed rank sum analysis to compare change in vitamin D and glutathione levels between baseline and 6-month and 12-month timepoints. We used Spearman correlation statistic to assess correlations between variables.

## RESULTS

### Study population

Among the 22 individuals screened 21 were enrolled (Stanford, n=18; Kennedy Krieger, n=3). The CONSORT Flow Diagram is presented in **Figure 1**. One participant was excluded because of an enhancing brain lesion on screening MRI. All participants completed all visits without loss to follow-up. The final two participant visits were completed remotely in March and June 2020 due to travel restrictions associated with a global coronavirus pandemic. Between October 2016 and December 2018, all participants (n=12) were started on a fixed dose of 2,000 IU of vitamin D daily. From January 2019 through end of recruitment in June 2019, remaining participants (n=9) were assigned to a weight-stratified dosing regimen with an initial 6-month dosing period of either 1,000 IU or 2,000 IU daily **(Figure 2)**.

**Figure 1:**
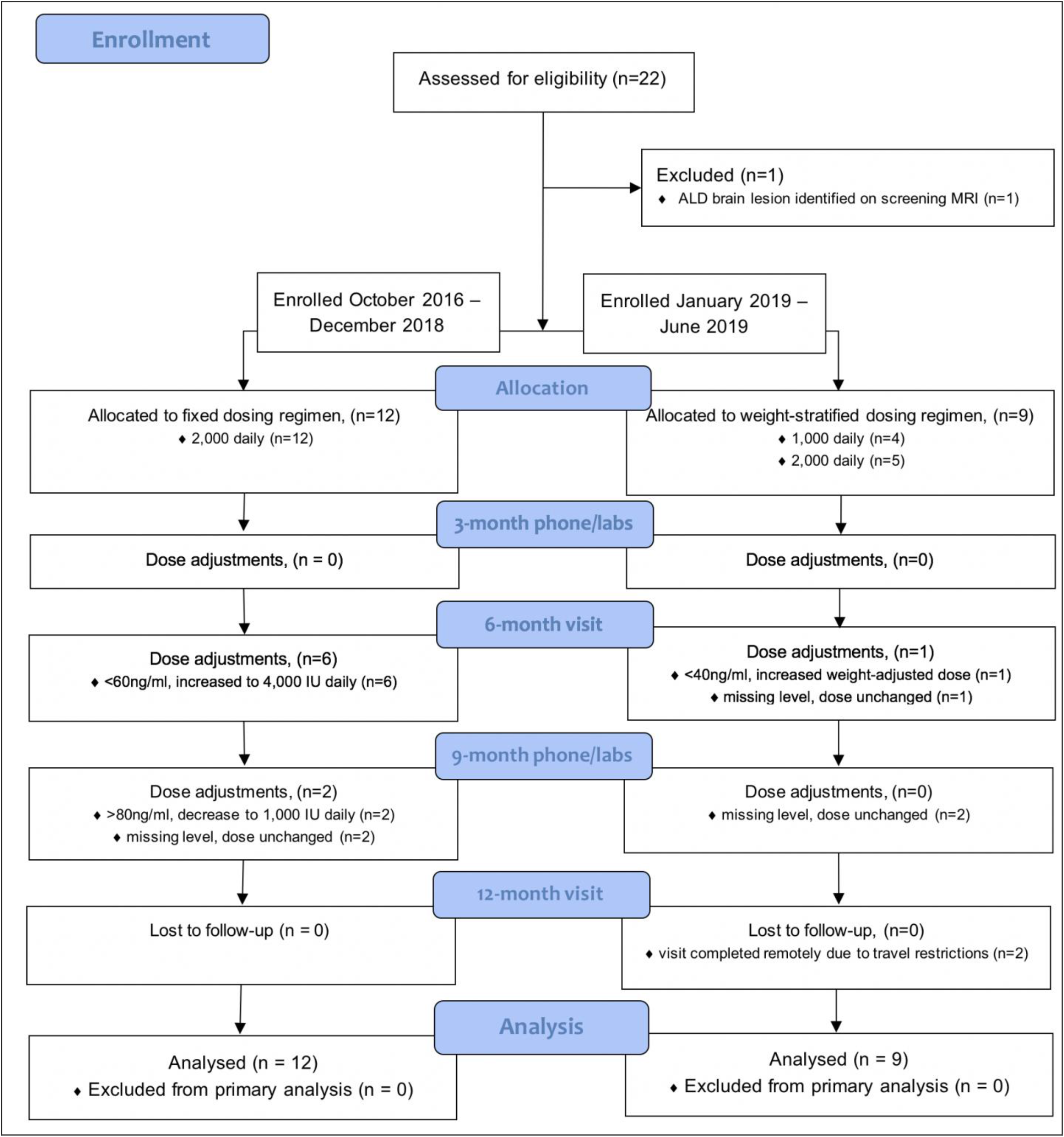
CONSORT Flow Diagram

**Figure 2:**
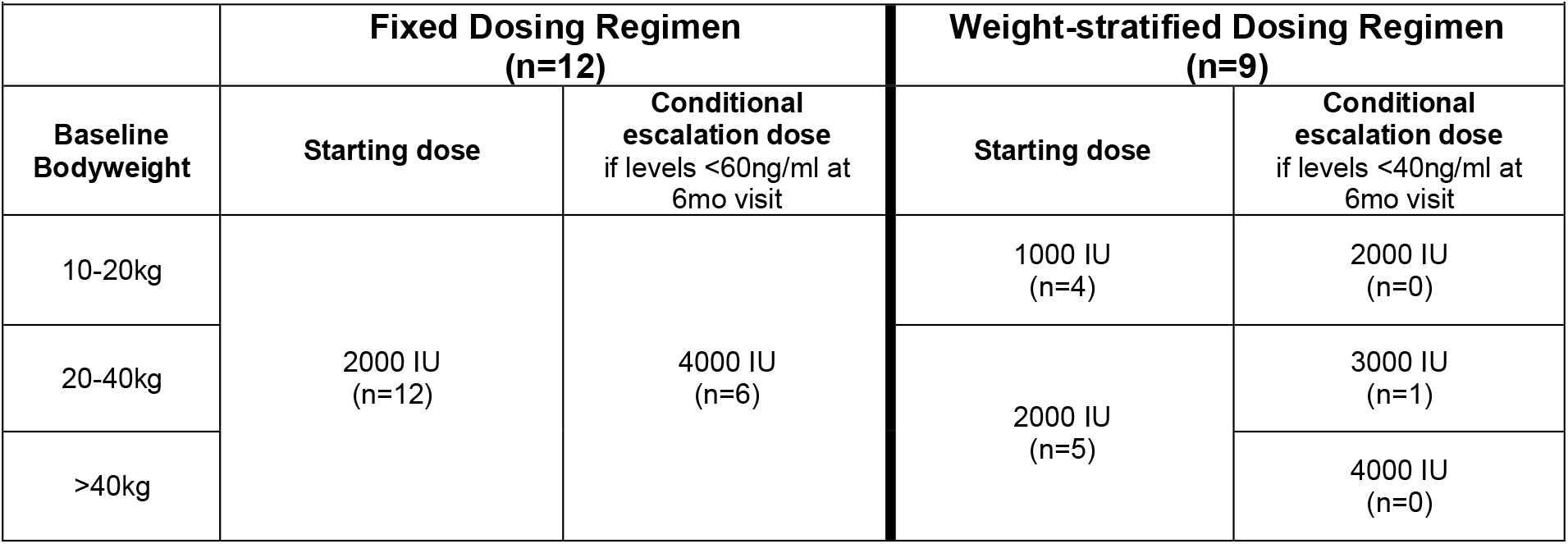
Dosing structure and participant tally for each dosing regimen

Demographic information and primary outcomes are summarized in **Table 1**. Overall, the median age (and interquartile range) was 6.7 years (9.2), the median baseline weight was 20 kg (29.5), and median baseline 25-hydroxyvitamin D level was 29 ng/ml (13). The fixed-dose group was somewhat younger (median age 6.4 vs 9 years), lighter (median weight 19.9kg vs 33.2kg) and had lower baseline vitamin D levels (median 28.5 vs 33.6ng/ml) than the weight-stratified group. Two patients in each dosing group had 25-hydroxyvitamin D levels greater than 40ng/ml at baseline with highest baseline level of 46ng/ml. Participants were primarily white, non-Hispanic (n = 15, 71%) which was similar across groups. Skin tones were similar between dosing groups. Weekly hours spent outdoors was stable across participants in fixed dose arm (**Figure 3**). The weight-based dosing arm, however, was affected by a surge in enrollment such that 8 of 9 participants enrolled in the first quarter of 2019, when sun exposure and vitamin D levels are at their annual nadir. This enrollment imbalance was reflected a peak in outdoor sun exposure (and to a less extent, 25-hydroxyvitamin D levels) at the mid-study point among weight-based participants.

**TABLE 1:** Demographics & Primary Outcomes.

|  | All participants<br>n=21 | By Dosing Regimen |  |
| --- | --- | --- | --- |
|  |  | Fixed<br>n=12 | Weight-stratified<br>n=9 |
| Demographics |  |  |  |
| Age, median (IQR), years | 6.7 (9.2) | 6.4 (8.3) | 9.0 (14.3) |
| Weight, median (IQR), kilograms | 20 (29.5) | 19.9 (21.4) | 33.2 (55.9) |
| Race/Ethnicity |  |  |  |
| White, non-Hispanic, n (%) | 15 (71) | 9 (75) | 6 (67) |
| Hispanic, n (%) | 5 | 4 | 1 |
| African American, n (%) | 1 | 0 | 1 |
| Asian, n (%) | 1 | 0 | 1 |
| More than one race, n (%) | 1 | 0 | 1 |
| Adrenal insufficiency at enrollment, n (%) | 12 (57) | 7 (58) | 5 (55) |
| Skin tone, median (IQR) | 3 (2) | 3 (1.5) | 3 (2.5) |
| Weekly time outdoors, median (IQR), hours | 17.5 (22.3) | 18.5 (19.6) | 8 (23) |
| Baseline 25-hydroxyvitamin D level, median (IQR), ng/ml | 29 (13) | 28.5 (11.1) | 33.6 (18.5) |
| Primary Outcomes |  |  |  |
| 6-month 25-hydroxyvitamin D level, median (IQR), ng/ml | 56.5 (14.8) | 58 (19.3) | 52.5 (15.5) |
| On target (40-80ng/ml), n (%) | 18 (86%) | 11 (92%) | 7 (78%) |
| Under target (<40ng/ml), n (%) | 3 (14%) | 1 (8%) | 2 (22%) |
| Over target (>80ng/ml), n (%) | 0 | 0 | 0 |
| 12-month 25-hydroxyvitamin D level, median (IQR), ng/ml | 61 (23.5) | 65.5 (15.8) | 44 (24.9) |
| On target (40-80ng/ml), n (%) | 14 (67%) | 8 (67%) | 6 (67%) |
| Under target (<40ng/ml), n (%) | 3 (14%) | 0 | 3 (33%) |
| Over target (>80ng/ml), n (%) | 4 (19%) | 4 (33%) | 0 |

**Figure 3:**
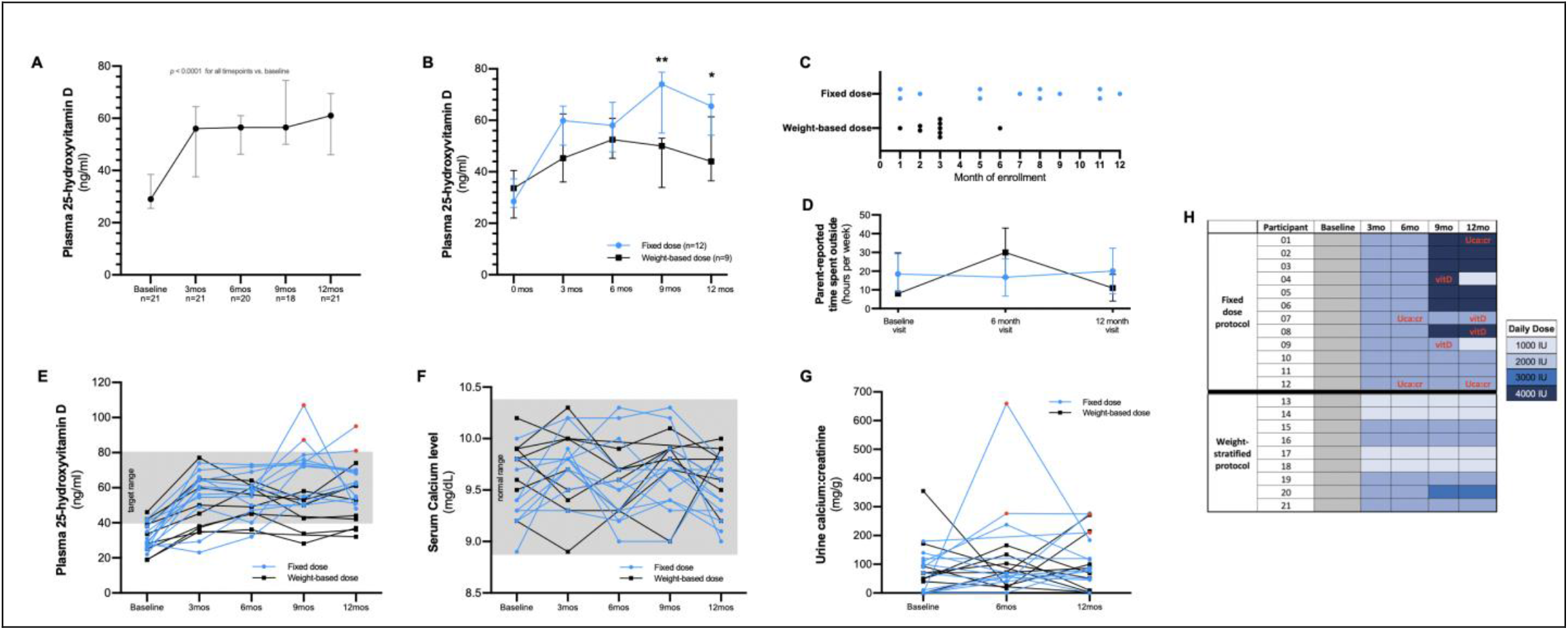
Plasma 25-hydoxyvitamin D levels were measured quarterly during the 12-month study period. Compared to baseline, plasma vitamin D levels were significantly higher at each timepoint (A). At 9-months and 12-months participants allocated to fixed dose of vitamin D achieved higher levels (B). Most participants in the weight-based cohort were enrolled in the first quarter of 2019 (C). This enrollment pattern yielded a corresponding imbalance in time spent outdoors for the weight-based cohort (D). Most participants achieved and maintained vitamin D levels within the specified target range of 40 – 80ng/ml, indicated by gray background; four patients exceeded the upper threshold (red dots) (E). All patients maintained normal serum calcium levels throughout the study (F). Normal ranges for urine calcium:creatinine levels vary by age, but were elevated in three patients at four time points (red dots) (G). Laboratory threshold violations are shown according by patient, regimen, and dose; red font indicates threshold violation for either plasma 25-hydroxy vitamin D level (vitD) or spot urine calcium:creatinine (Uca:cr), (H). No laboratory threshold violations were observed among participants in the weight-stratified dosing regimen.

### Primary outcome

At 6 months, 92% (n = 11) in the fixed-dose group and 78% (n = 7) in the weight-stratified group achieved target 25-hydroxyvitamin D levels (40-80 ng/ml). Half of participants in the fixed-dose group (n=6) qualified for dose escalation (level <60ng/ml); one participant in the weight-stratified group qualified for dose escalation (level <40ng/ml). One participant in the weight-stratified group did not complete a 6-month vitamin D level and remained on starting dose. At 12 months, 67% of both groups achieved target levels without a triggered dose reduction (n = 8 in fixed-dose; n = 6 in weight-stratified). **Figure 3** displays quarterly measures of plasma 25-hydroxyvitamin D levels during the 12-month study period. Overall, median levels were similar between groups except at months 9 and 12 when plasma 25-hydroxyvitamin D levels were higher in the fixed-dose group.

### Safety and adverse events

Adverse events (AEs) occurring over the 12-month study period are summarized in **Table 2**. Safety laboratory levels (i.e. plasma 25-hydroxyvitamin D, serum calcium, and random urinary calcium:creatinine) are summarized in **Figure 3**. Both pre-defined and general adverse events occurred more frequently among participants in the fixed-dose arm. Among all safety laboratory measures (n=202) of plasma 25-hydroxyvitamin D, serum calcium, and urine calcium:creatinine, we observed 8 laboratory elevations in 6 participants, all of whom were assigned to the fixed-dose regimen (**Figure 3**). Four participants had 25-hydroxyvitamin D levels above pre-defined safety threshold of 80ng/ml with two events among participants receiving a daily dose of 2,000 IU daily and two receiving 4,000 IU daily (**Figure 3C, 3F**). Similarly, transient elevations in urine calcium:creatinine were also observed in the fixed-dose arm (n=2 at 2,000 IU; n=1 at 4,000 IU) daily; no persistent elevations in urine calcium:creatinine were observed in any participants. Serum calcium levels remained in the normal range for all participants. Brain MRI and neurologic assessments also remained normal in all participants.

**TABLE 2:**
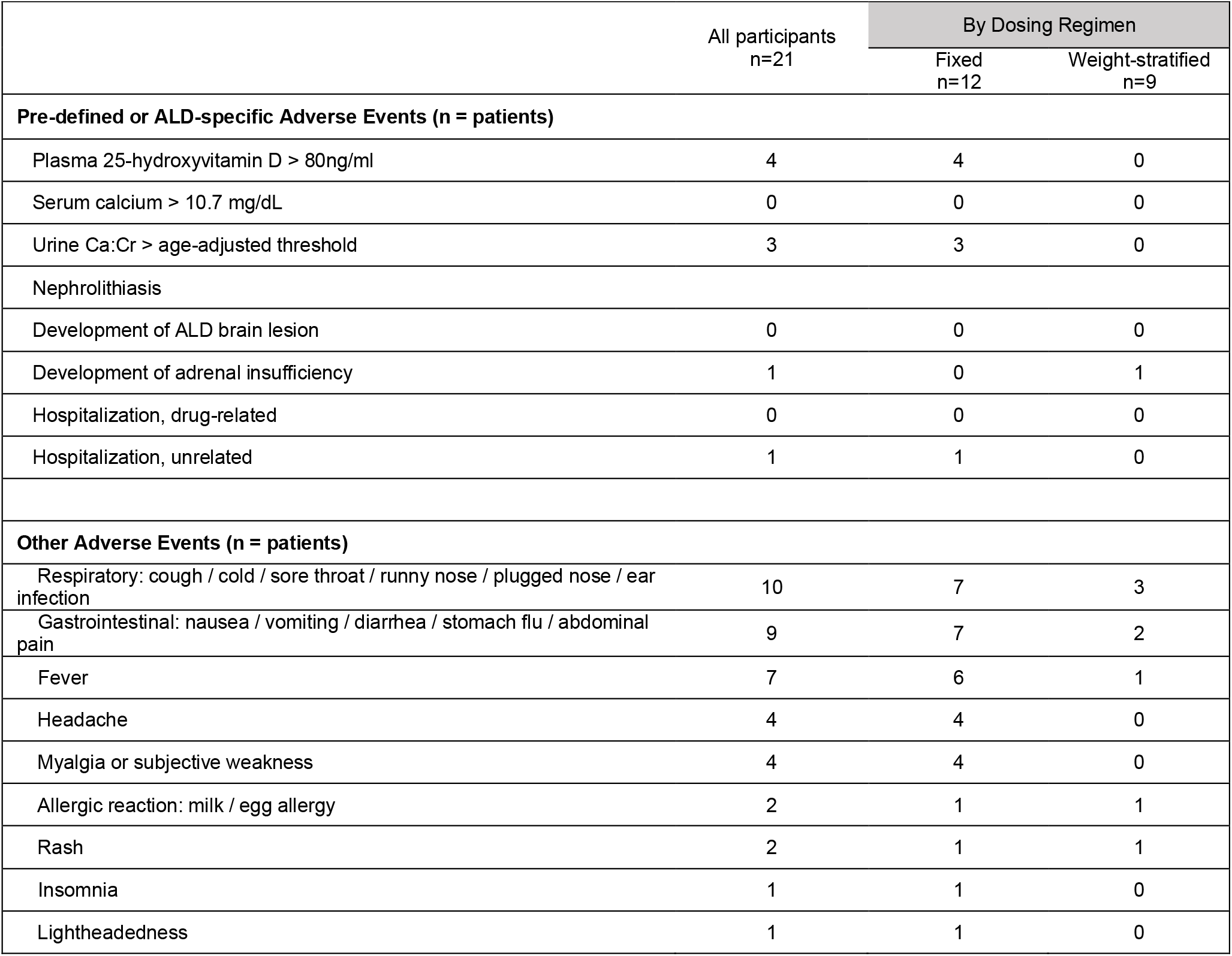
Frequency of adverse events

|  | All participants<br>n=21 | By Dosing Regimen |  |
| --- | --- | --- | --- |
|  |  | Fixed<br>n=12 | Weight-stratified<br>n=9 |
| Pre-defined or ALD-specific Adverse Events (n = patients) |  |  |  |
| Plasma 25-hydroxyvitamin D > 80ng/ml | 4 | 4 | 0 |
| Serum calcium > 10.7 mg/dL | 0 | 0 | 0 |
| Urine Ca:Cr > age-adjusted threshold | 3 | 3 | 0 |
| Nephrolithiasis |  |  |  |
| Development of ALD brain lesion | 0 | 0 | 0 |
| Development of adrenal insufficiency | 1 | 0 | 1 |
| Hospitalization, drug-related | 0 | 0 | 0 |
| Hospitalization, unrelated | 1 | 1 | 0 |
| Other Adverse Events (n = patients) |  |  |  |
| Respiratory: cough / cold / sore throat / runny nose / plugged nose / ear infection | 10 | 7 | 3 |
| Gastrointestinal: nausea / vomiting / diarrhea / stomach flu / abdominal pain | 9 | 7 | 2 |
| Fever | 7 | 6 | 1 |
| Headache | 4 | 4 | 0 |
| Myalgia or subjective weakness | 4 | 4 | 0 |
| Allergic reaction: milk / egg allergy | 2 | 1 | 1 |
| Rash | 2 | 1 | 1 |
| Insomnia | 1 | 1 | 0 |
| Lightheadedness | 1 | 1 | 0 |

One participant, assigned to the fixed-dose regimen, was hospitalized for an asthma attack which was deemed unrelated to study drug. One participant, assigned to the weight-stratified regimen, developed adrenal insufficiency. Among the other AEs, participants in the fixed-dose group experienced more fever, headaches, myalgia/subjective weakness, respiratory and gastrointestinal AEs; however, this may be due to differences in study time period (overlap with flu season) and/or differences in age between the two dosing groups. Other AEs experienced were allergic reaction, rash, insomnia, and lightheadedness.

### Glutathione levels in brain and blood

Spectroscopy was acquired at 57 of 63 study timepoints. Six acquisitions could not be completed due to travel restrictions (n=4) or software error (n=2). Among the 114 acquired spectra, two from the occipital white matter and 13 from the frontal white matter did not meet quality threshold and were excluded. Voxels in the frontal white matter were adjacent to sinuses with poor B_0_ inhomogeneity.

Brain glutathione levels correlated across frontal and occipital white matter regions within subjects at each visit suggesting good internal validity (**Figure 4C**; Spearman r=0.43; p=0.005). Brain glutathione levels in both the occipital white matter (**Figure 4D**) and frontal white matter (**Figure 4E**) were higher at 12-months compared with baseline. Brain glutathione levels did not correlate with plasma vitamin D levels in either brain region (**Figure 4G, 4H**).

**Figure 4:**
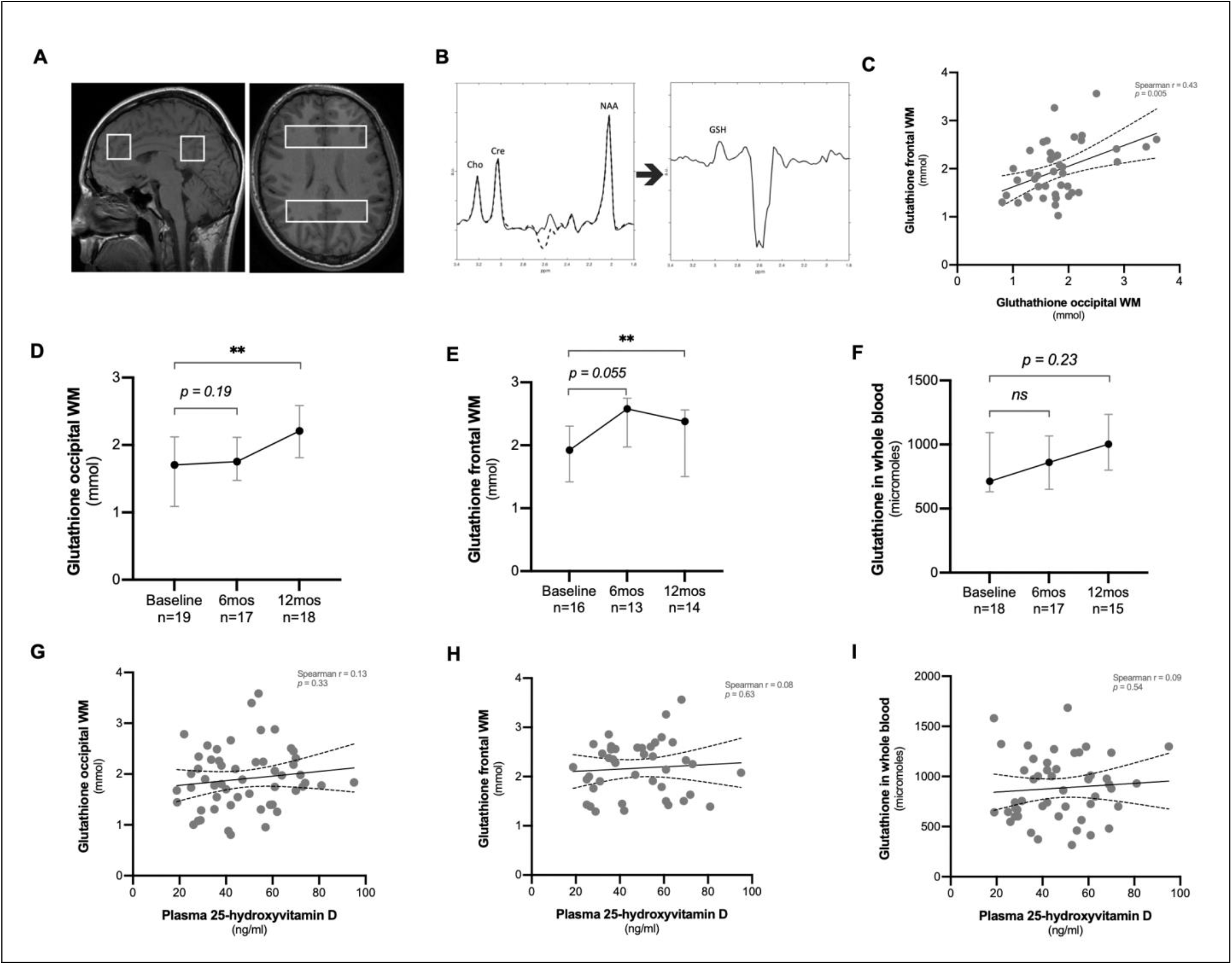
Glutathione levels were measured in the blood and brain at baseline and again after 6 and 12 months of vitamin D supplementation. We used magnetic resonance spectroscopy to measure brain glutathione levels in the frontal and occipital white matter where cerebral ALD lesions typically begin (A). MEGA-PRESS acquired spectra with editing on (dotted line) and editing off (solid line) are shown alongside the edited spectrum which delineates the GSH peak at 2.95 ppm (B). Measures of glutathione in frontal white matter correlated with measures in occipital white matter (C). Glutathione levels increased between baseline and 12 months in occipital white matter and frontal white matter, but not in plasma (D, E). Glutathione levels did not increase significantly in plasma (F). Glutathione levels in the brain blood did not correlate with plasma vitamin D levels (G, H, I). Figures C, G, H, I display all available data. In Figures D, E, F median (dot) and interquartile range (bars) are shown for all participants for whom a baseline measure was available; Wilcoxon matched-pairs signed rank analysis was used for all comparisons between baseline and 6-month and 12-month timepoints.

Plasma samples for glutathione analysis were available from 49 timepoints. Glutathione levels were not significantly different between baseline and 6-month and 12-month timepoints and did not correlate with vitamin D levels. (**Figure 4F, 4I**).

## DISCUSSION

In this phase I study of oral vitamin D3 supplementation in 21 ALD boys aged 19mos to 22 years, a weight-stratified dosing regimen achieved similar rates of 25-hydroxyvitamin D target levels as a fixed-dose regimen and had fewer safety laboratory elevations and fewer adverse events overall. No participants experienced clinical adverse events attributable to vitamin D. Glutathione levels in the frontal and occipital white matter increased during the 12-month study period.

Vitamin D dosing regimens in children have historically targeted levels >20ng/ml to protect against rickets with comparatively little focus on achieving higher levels that are more commonly sought for the treatment of multiple sclerosis^11-14, 17, 25, 26^. Our study suggests that our weight-based dosing regimen was safe and clinically well-tolerated. The unexpected peak in plasma 25-hydroxyvitamin D levels at the mid-study timepoint was in the weight-based regimen was likely due to enrollment surge in the 1^st^ quarter of 2019 that produced a mid-study peak in sun exposure corresponding with summer and early when sun exposure and 25-hydroxyvitamin D levels are highest. Several participants assigned to our fixed-dose regimen achieved vitamin D levels above our target threshold although none manifested hypercalcemia or associated signs and symptoms. The higher levels in the fixed dosing regimen resulted from higher vitamin D dose exposures.

The fixed-dose group recorded more laboratory deviations and non-specific adverse events than the weight-stratified group. The adverse events reported in both groups were non-specific and common among children (e.g., abdominal or respiratory symptoms). Although none of these reported adverse events were considered attributable to vitamin D supplementation, it is possible that higher doses of vitamin D may predispose to mild, but common and nonspecific ailments. One participant in the fixed-dose group was hospitalized for an asthma attack, which was not attributable to study drug. Although vitamin D supplementation has a demonstrated record in reducing the risk of respiratory infections and asthma attacks, this effect could be dose-dependent^27-30^. Urine calcium:creatinine levels were elevated in 3 individuals receiving the fixed-dose regimen, but were asymptomatic and self-resolved without intervention.

Symptoms of vitamin D toxicity, which include vomiting, weakness, and anorexia, overlap with symptoms of adrenal insufficiency. However, vitamin D toxicity is caused by hypercalcemia which distinguishes it from adrenal insufficiency and was not observed in any of our study participants. Although no participants reported adrenal crises during the study period, one participant, in the weight-stratified group, developed ALD-related adrenal insufficiency that was observed during routine laboratory surveillance.

Glutathione is a ubiquitous antioxidant whose deficiency predisposes to cellular injury and death in the brain and immune system^31-33^. Low glutathione levels have been repeatedly observed in ALD boys and men^34-36^. Median levels of glutathione in the brain increased over the 12-month study period. Median levels of glutathione in whole blood were higher at 6 and 12-months but were not statistical different than baseline. We did not observe a correlation between plasma 25-hydroxyvitamin D and plasma or brain glutathione levels, which could be explained by our small sample size and/or the non-linear pharmacodynamics of 25-hydroxyvitamin D biology^11, 27, 29^. The absence of a placebo control limits causal inference between vitamin D and glutathione in our sample; however, our observations are consistent with a recent and growing body of randomized placebo-controlled clinical trials linking vitamin D supplementation with increased glutathione levels in blood.^18-21^ Our study is the first, to our knowledge, to examine this phenomenon in a pediatric population and the first to assess the effects of vitamin D supplementation on brain glutathione.

Among our study’s limitations were our small sample size and lack of a placebo control. Our 12-month study was also not designed to assess the potential for long-term risks associated with high-dose vitamin D supplementation. Although previous studies suggest spectroscopy results can be reliably reproduced across clinical sites and machine vendors, technical differences may persist across sites^37^. We did not account for the possibility of age-related changes in brain glutathione during childhood although significant changes over a one-year period seem unlikely. Finally, our study was not powered to assess vitamin D’s ability to mitigate the onset of cerebral ALD lesions, although none were observed.

Future studies of vitamin D supplementation in ALD should consider implementing a weight-stratified dosing regimen and focus on vitamin D’s potential efficacy in preventing the formation of brain lesions. Such a study should also allow the opportunity to further study vitamin D’s impact on adrenal function in ALD. A prevention trial in ALD will present a design challenge due to the long period of follow-up required and known challenges associated with randomization in vitamin D studies, particularly when the target outcome is severe^38^.

In summary, we find that daily dosing of vitamin D3 supplementation, particularly when stratified by weight, can safely achieve plasma 25-hydroxyvitamin D levels between 40-80ng/ml in most boys with ALD. Our findings also implicate brain glutathione as a candidate biomarker for vitamin D supplementation in ALD, although a causal association was not established in this study. These findings are of interest to future studies assessing vitamin D’s role in preventing brain lesions in ALD.

## Data Availability

All data produced in the present study are available upon reasonable request to the authors

https://clinicaltrials.gov/ct2/show/NCT02595489

## Acknowledgements

Special thanks to the study participants and their families; thanks also to Julianne Jorgensen, Sweta Patnaik, Simone Schubert, and Fe Gibbons who supported study logistics.

